# The impact of differentiated service delivery (DSD) on retention in care and viral suppression in South Africa: A target trial emulation of routine care data

**DOI:** 10.1101/2024.10.11.24315285

**Authors:** Amy N. Huber, Lise Jamieson, Matthew P. Fox, Musa Manganye, Lufuno Malala, Thato Chidarikire, Nthabiseng Khoza, Brooke E. Nichols, Sydney Rosen, Sophie Pascoe

**Affiliations:** Health Economics and Epidemiology Research Office, Faculty of Health Sciences, University of the Witwatersrand, Johannesburg, South Africa; The South African Department of Science and Innovation/National Research Foundation Centre of Excellence in Epidemiological Modelling and Analysis (SACEMA), Stellenbosch University, Stellenbosch, Republic of South Africa; Department of Global Health, Boston University School of Public Health, Boston, MA, USA; National Department of Health, Pretoria, South Africa; World Health Organization; Department of Global Health, Amsterdam Institute for Global Health and Development, Amsterdam UMC, University of Amsterdam, Amsterdam, Netherlands

**Keywords:** South Africa, HIV, antiretroviral therapy, differentiated service delivery, outcomes

## Abstract

**Introduction:** Replacing conventional, facility-based HIV treatment with less intensive differentiated service delivery (DSD) models could benefit DSD clients and the health system, but its value depends on maintaining or improving clinical outcomes. We compared retention and viral suppression between antiretroviral therapy (ART) clients enrolled in DSD models to those eligible for but not enrolled in DSD models in South Africa.

**Methods:** We applied a target trial emulation (TTE) methodology to data from South Africa’s electronic medical record system (TIER.Net) for 18 clinics across 3 provinces and estimated retention in care (attended clinic visit within 12 months) and viral suppression (<400 copies/ml^3^) at 12, 24, and 36 months after follow-up start date, defined as DSD enrollment date for the intervention arm and the first trial enrollment period clinic visit for the comparison arm. Clients were eligible for DSD models if they were ≥18 years old, on ART ≥12 months, and had two suppressed viral load (VL) measurements, per prevailing national guidelines. For the TTE, we designated eight 6-month target trial enrolment periods between 1 July 2017 and 1 July 2021. For each period, we estimated the risk differences for retention in care and viral suppression by comparing those enrolled in DSD models to those not enrolled, using a Poisson distribution with an identity link function. We report adjusted and unadjusted risk differences for clients enrolled in DSD models and for DSD-eligible clients not enrolled in a DSD model.

**Results and discussion:** 49,595 unique individuals were eligible for DSD enrolment over eight target trials, contributing to a total of 148,943 trial-clients, of whom 17% (25,775) were enrolled in DSD models. The pooled adjusted risk difference for retention in care between clients enrolled in DSD and those not enrolled in DSD was 3.2% (95% confidence interval (CI) 1.6%; 4.7%) at 12 months, 4.2% (2.4%; 6.0%) at 24 months, and 4.4% (2.0%; 6.8%) at 36 months. For viral suppression, the adjusted risk difference comparing DSD to non-DSD was estimated to be 1.4% (-0.5%; 3.2%) at 12 months, 1.7% (-0.5%; 4.0%) at 24 months, and 1.4% (- 0.6%; 4.4%) at 36 months. Results remained consistent across target trials. Clients who were younger, lived in urban settings, or had less ART experience at trial enrolment had lower retention.

**Conclusions:** Clients enrolled in DSD models in South Africa had slightly better retention in care and similar viral suppression to those who were eligible for but not enrolled in DSD. With better or equivalent outcomes, DSD models can be assessed on the basis of non-clinic costs and benefits, such as changes in quality of care and resource utilization.

**Registration:** Clinicaltrials.gov NCT04149782.

## Introduction

In South Africa, as in other countries in sub-Saharan Africa (SSA), the use of differentiated service delivery (DSD) models for HIV treatment is expanding rapidly [1,2]. For eligible antiretroviral therapy (ART) clients, DSD models adapt the characteristics of service delivery, such as location and frequency of healthcare system interactions, to meet the needs of different client groups. This approach is expected to sustain clients’ positive clinical outcomes, increase satisfaction with their care, and increase the efficiency of healthcare systems by decongesting clinics, reducing pressure on healthcare providers, and simplifying procedures[3,4]. Most DSD models are limited to “established” or stable ART clients who have been on treatment for at least 6 months, have documented viral load suppression, and do not have uncontrolled co-morbid conditions or HIV-related symptoms[5]. The proportion of established clients enrolled in any DSD model, rather than remaining in conventional (non-differentiated) care, varies widely by country and facility, based on guidelines, resources, and provider and client preferences, among other factors[6].

South Africa, home to the world’s largest HIV treatment program[7], offers three main less intensive models for ART clients who are established on treatment and have documented viral suppression: facility-based medication pickup points, external (out-of-facility) medication pickup points, and adherence clubs. (In South Africa, these models are referred to as Differentiated Models of Care, or DMOC and fall within the category of Repeat Prescription Collection strategies or RPCs.) The three models for established ART clients are supported by the Central Chronic Medicine Dispensing and Distribution (CCMDD) programme, the Central Dispensing Unit (CDU), or facility-based pharmacies. They provide centralized medication packaging and delivery to clients, with options for pickup at either facility pickup points within healthcare facilities or at external pickup points adjacent to facilities or within communities[7–10]. Other DSD models, such as medication lockers and home delivery, have also been implemented in South Africa, but on a much smaller scale.

Several randomized trials and observational studies have reported the outcomes of clients enrolled in South Africa’s less intensive models [11–13] and in others outside South Africa[14] and found high rates of retention in care and viral suppression after one or more years of model exposure, an unsurprising result given that retention and suppression are also both eligibility criteria for model enrollment. Few studies[15–17], however, have compared the outcomes of clients actively enrolled in DSD models to those who are eligible for DSD models but remain in conventional care. It is these eligible-but-not-enrolled clients who comprise the relevant comparison group.

The absence of evidence about how clients’ outcomes in DSD models compare to those of similar clients remaining in conventional care hampers decisions about whether to expand differentiated service delivery programs and how to update eligibility criteria. To help fill that gap, we assessed the effectiveness of DSD by comparing clients on DSD to those eligible for but not on DSD on outcomes of retention in care and viral suppression at 12, 24, and 36 months using routinely collected electronic medical record data.

## Methods

Using a target trial emulation approach, we analyzed routinely collected medical records from a sample of public sector facilities. We compared the 12-, 24- and 36-month retention in care and viral suppression outcomes of DSD clients to those eligible but not enrolled in DSD in South Africa at primary health clinics between 1 July 2017 and 1 July 2021.

### Models of care

As noted above, DSD-eligible ART clients were either enrolled in one of three less intensive differentiated models of care in use during our analysis – primarily facility-based medication pickup and external medication pickup, along with adherence clubs for a smaller number of clients – or in conventional (non-differentiated) care. Enrollment decisions were presumably based on client eligibility, provider preference, the availability of each model at any specific healthcare facility, and the facility’s access to enrolment procedures (e.g. the CCMDD database). Features of the four models represented in our analysis are described in Table 1. We were not able to distinguish between these in the TIER.Net data and therefore could only evaluate the impact of all DSD models compared to conventional care, rather than comparing DSD models with one another. The 2016 South African Adherence Guidelines were used to define DSD eligibility and viral suppression in this analysis, as the majority of the enrollment period fell within their period of coverage [18]. During the study period, eligibility for DSD enrollment required clients to be non-pregnant adults with a minimum of 12 months of experience on ART and at least two documented suppressed viral loads (<400 copies/ml). We note that under current guidelines, the VL threshold for suppression is 50 c/mL and 3 month-dispensing is recommended.

**Table 1.**
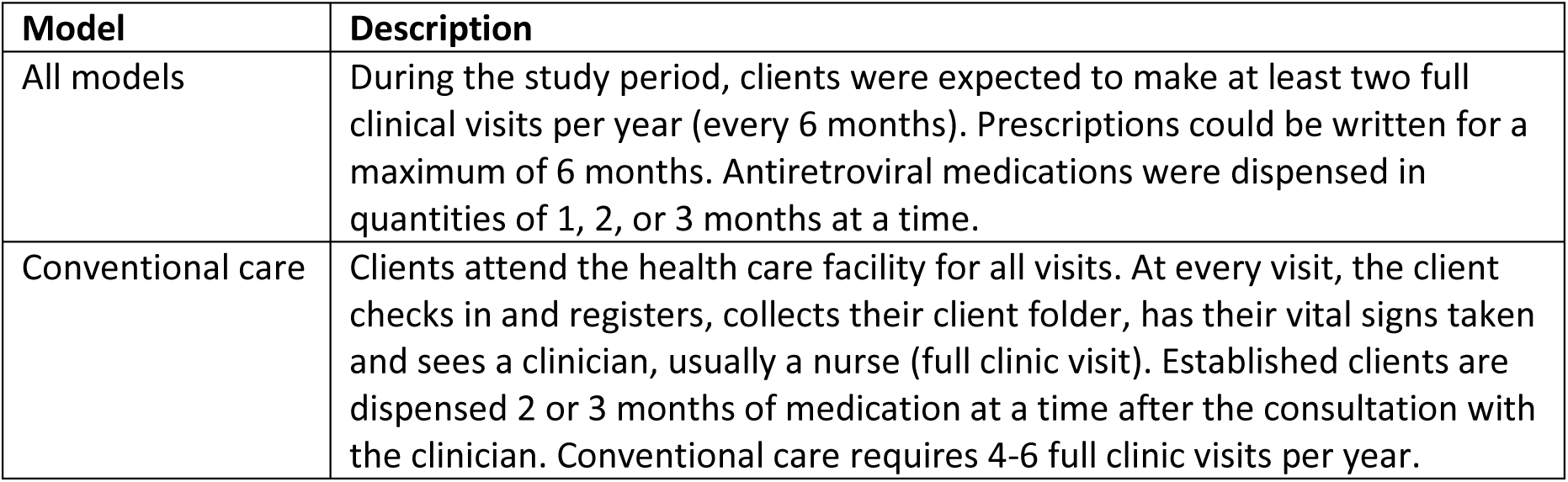
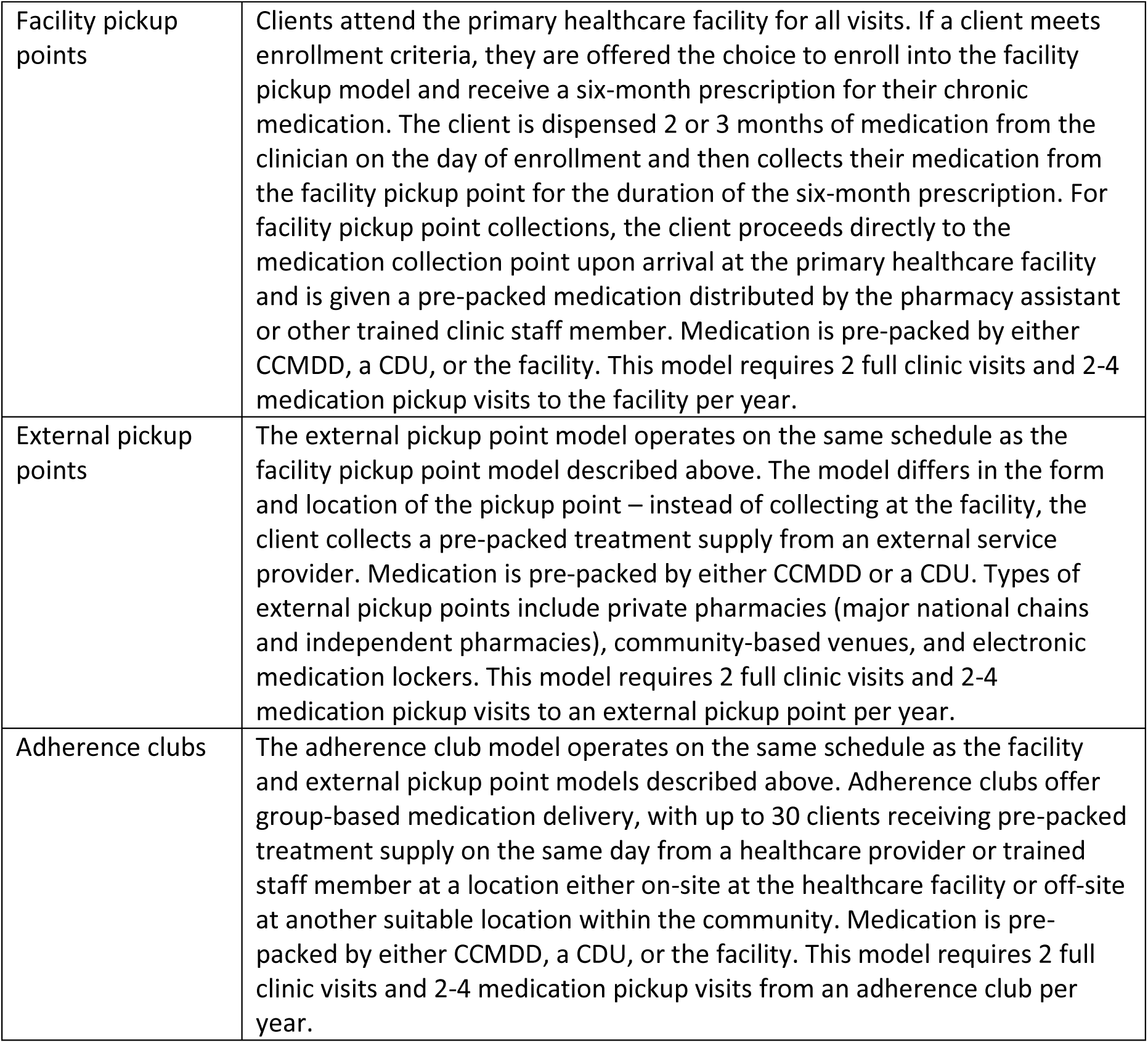
Description of widely used models of HIV treatment delivery in South Africa.

### Study sites and population

Data were drawn from 24 public-sector health facilities in four districts in three provinces of South Africa (6 clinics in each district-West Rand and Ekurhuleni in Gauteng Province, King Cetshwayo in KwaZulu-Natal Province, and Ehlanzeni in Mpumalanga Province). We purposively chose a mix of high- and medium-volume facilities in urban and rural settings. All sites included offered at least one DSD model at the time of data collection had at least 1,000 clients on ART and utilized South Africa’s electronic medical record system, TIER.Net[19]. The data extracted from the study sites spanned the period from July 2017 to June 2023, depending on the district, and in 2021 the approximate proportion of ART clients enrolled in DSD models ranged between 24% and 81%, with KwaZulu-Natal reporting the highest proportion of clients enrolled. (Supplementary Table S1).

The TIER.Net system[20] is a facility-based electronic register. Each client’s health information is recorded in a paper file at the point of care and then entered into TIER.Net by an on-site data capturer. Data are collected prospectively, and inactive records are not deleted. The system allows for capture of age, gender, HIV test date, ART start date, visit date, ART medication prescribed, and months of ART dispensed for all clinic interactions, date and result of HIV-related laboratory tests, and the outcome date and status for clients who have died, transferred, or become disengaged from care. A DSD model enrollment variable was introduced in 2016, but with three limitations for DSD program monitoring. First, out-of-facility medication collection from external pickup points and community-based adherence clubs is not captured in TIER.Net—only actual clinic visits are recorded. Second, before June 2020, the DSD enrollment variable did not distinguish among separate DSD models, but only the client’s enrollment in any non-conventional model. And third, unofficial (silent) transfers of clients between facilities are not captured in TIER.Net, creating a potentially large discrepancy between facility-level and overall ART program loss to follow-up [21]. To address these limitations, we grouped all DSD models together, included a 12-month window for follow-up visit measurement, and reported facility-level retention.

### Target trial emulation

Target trial emulation is a methodology that simulates a hypothetical randomized clinical trial and can be applied to observational data in a way that assists in estimating causal effects[22]. It also helps to prevent immortal time bias, whereby a person can accrue person time in the study when they cannot logically experience the outcome. Immortal time bias often occurs when the beginning of follow-up time and intervention exposure are not aligned[23]. Using this methodology, we clearly described the hypothetical trial we would like to have conducted and then attempted to mimic that trial as closely as possible using observational data, potentially reducing bias in our assessment of the effect of DSD on treatment outcomes.

Table 2 outlines our hypothetical target trial and the choices we made in our observational study to best emulate that trial. Both the target trial and the emulation sought to estimate the impact of enrollment in differentiated service delivery (DSD) models on retention in care and viral load suppression at 12-, 24- and 36-months after DSD eligibility. To do this, we used in both the trial and the emulation the DSD eligibility criteria in the South African guidelines [21] in effect at the time of the study [18]. In addition, we included in the emulation additional trial eligibility criteria in line with the conduct of our hypothetical trial: 1) documentation of a recent clinic visit, within 6 months before the trial, and 2) no prior DSD enrollment recorded (DSD naïve at study enrollment).

**Table 2.**
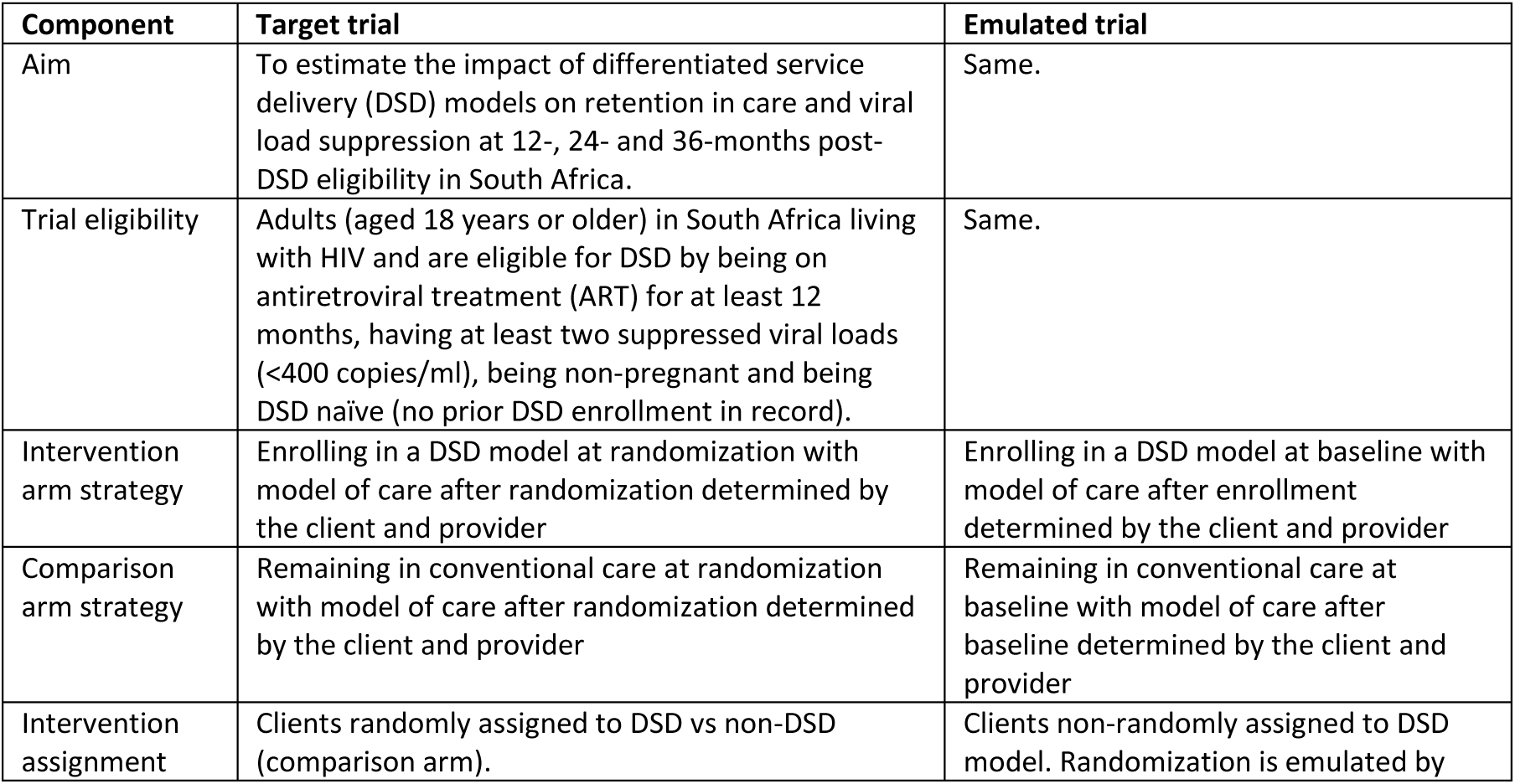
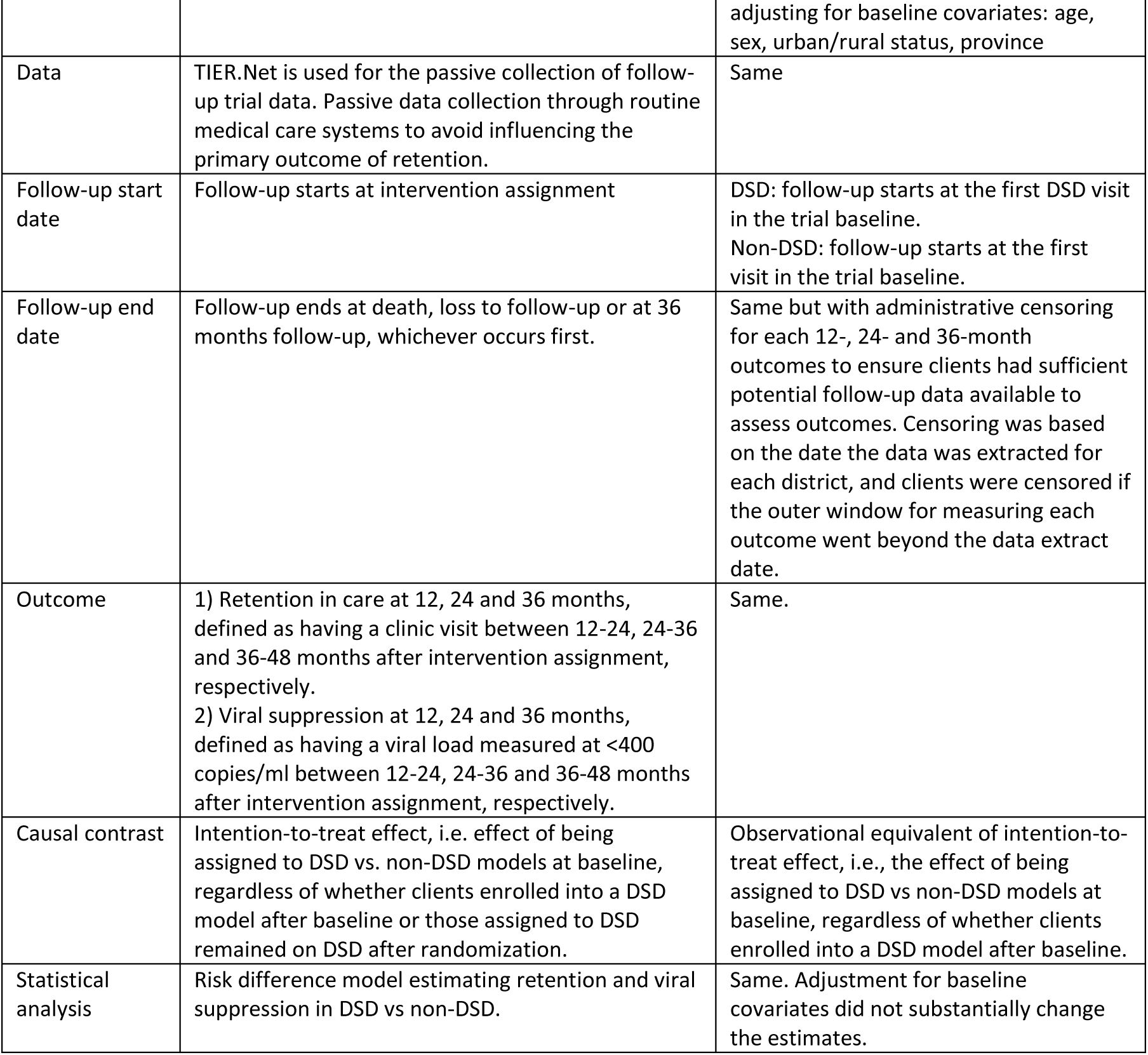
Target trial emulation specification of differentiated service delivery (DSD) models care.

Our target trial intervention was enrollment in a DSD model. After target trial randomization, participants could return to standard care at the discretion of the client and the provider. We mimicked this in our emulation by defining the intervention to be enrollment in DSD at baseline. After DSD enrollment, participants could return to regular standard care, but we would still analyze them as if they were in the DSD arm as per intention-to-treat, as would have been done in the target trial. The comparison arm for our target trial was the participants randomized to not immediately enroll in a DSD model. As would happen in routine care, however, comparison arm participants would be allowed to enroll in a DSD model at a later time point after the enrollment period, should they be offered the opportunity and accept it. This effectively alters our study question to comparing the effect of enrolling in DSD immediately to the effect of a delayed opportunity for DSD enrollment, with some comparison arm participants later enrolling in DSD and some never doing so. To mimic this in our emulated trial, we assigned anyone within the baseline enrollment period who did not enroll in DSD to the comparison arm, noting that after baseline, they had regular standard care, which in some cases could involve later enrolling in DSD.

### Development of multiple trial emulations

For this analysis, we chose to emulate a series of eight target trials, each encompassing a 6-calendar month DSD enrollment period in our data set as our baseline, between 1 July 2017 and 1 Jul 2021 (e.g. 1 July 2017 to 31 December 2017, 1 January 2018 to 30 June 2018, etc.). This approach maximized the value of our multi-year data set within the constraints of the target trial methodology. Had we conducted only one emulated trial limited to the first six-month calendar period for which we had data (July to December 2017), all participants could readily have been assigned to either intervention or comparison arm status, but this approach would have excluded all the individuals in the data set who enrolled in or became eligible for DSD enrollment after December 2017. Alternatively, we could have conducted only one emulated trial incorporating all the data in our data set (July 2017 to June 2021), in which all clients who enrolled in DSD later in the study period would have been assigned to the DSD arm. As those clients would, by definition, have survived longer on ART before enrolling in DSD, however, and longer duration on ART is associated with better retention and suppression outcomes, this could create bias.

Emulating multiple, time-limited trials offers one solution to the problem described above. Within each of the eight target trial periods we assessed eligibility for participation in the trial and DSD models for individuals in the database at the start of each six-month trial (Figure 1). All those who met the criteria were included in the new trial, and individual clients could thus be included in the comparison arm of multiple trials. Participants eligible for each six-month calendar period were then assigned to the intervention or comparison arm. Because being DSD-naïve is a trial eligibility criterion, anyone who had enrolled in DSD during a prior six-month period and been analyzed in a previous emulated trial would no longer be eligible for subsequent trials. A participant who had not yet enrolled in DSD in a prior period could meet the study eligibility criteria for subsequent trials, however, along with anyone who became newly eligible for DSD based on national guidelines. As a result, any individual could be enrolled in one or more trials as a comparison arm participant only; in one or more trials as a comparison arm participant and one as an intervention participant; or in just one trial as an intervention participant.

**Figure 1:**
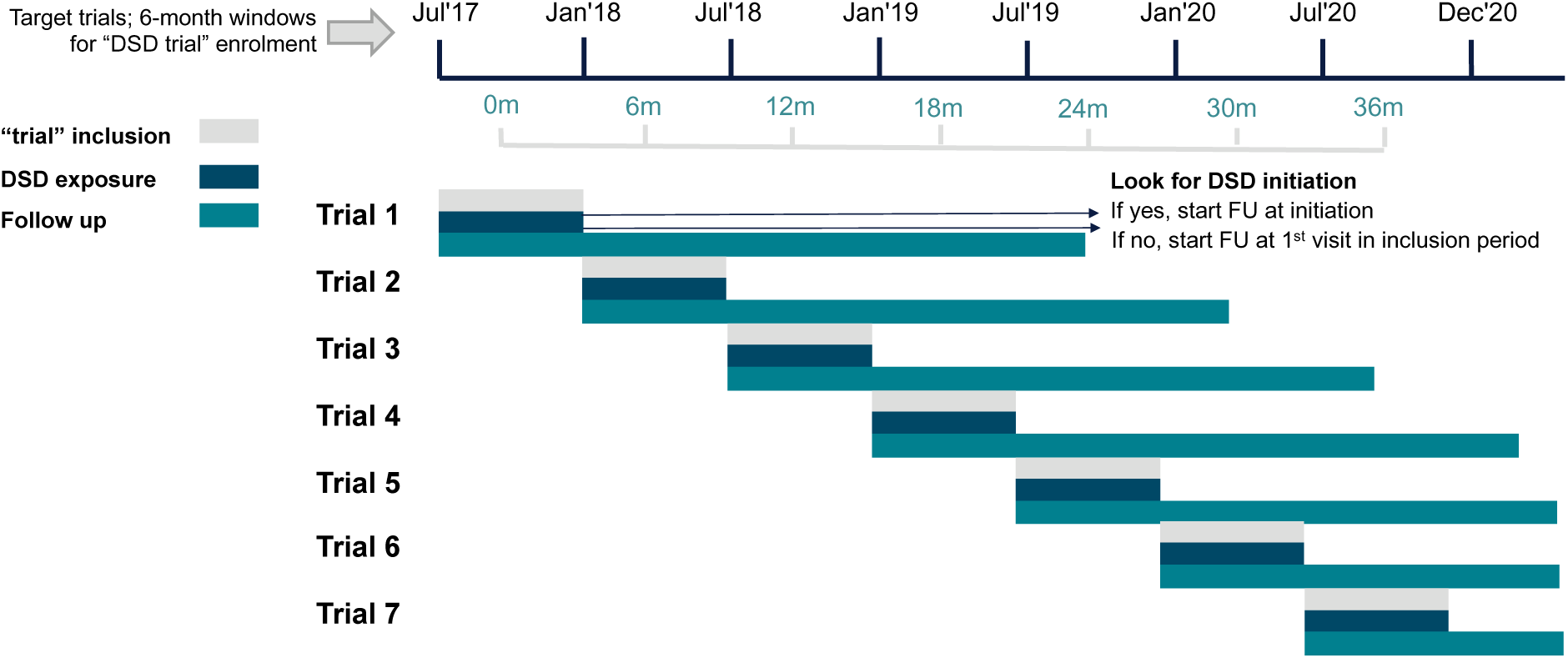
Summary of the eight target trial periods.

### Outcomes

We defined two outcomes for this analysis, retention in care and viral suppression, each assessed at 12, 24, and 36 months after the start date of follow-up as specified in Table 2. Retention in care was defined as having had a recorded clinic visit at the same clinic facility at 12, 24, or 36 months after the start date, with the start date defined as the DSD enrolment date for the intervention arm and the first clinic visit attended during the trial enrolment period for the comparison arm. Viral suppression (<400 copies/mL) was also assessed at these time points but could only be reported for those with viral loads measured (i.e. our denominator only includes those who had documented viral load results in their medical records); participants missing viral load measures were excluded from the analysis of this second outcome. It is important to note that because our data source, TIER.Net, does not capture unreported (“silent”) transfers between facilities, those who informally transfer appear to have disengaged from care. Retention in this study thus refers to retention at the same facility, not retention across the HIV treatment program.

For both retention in care and viral suppression outcomes, we allowed for a wide 12-month window after the endpoint to recognize outcomes. Clients were considered retained in care for 12 months, for example, if they had a clinic visit any time between 12 and 24 months after DSD enrolment, and their viral load outcome for the 12-month endpoint reflected laboratory tests conducted between 12 and 24 months after DSD enrollment. These relatively long (12-month) windows were chosen to accommodate local practice during the study period, which included the COVID-19 pandemic when South Africa temporarily allowed 12-month prescriptions of ART medications. A 12-month window for recording viral suppression was necessary because viral load tests are recommended and typically conducted only every 12 months after the first year on ART. We used the earliest viral load result within the 12-month window after the specified endpoint as the outcome measure. We also extracted from TIER.Net individual characteristics such as age, gender, time on ART, WHO stage, and CD4 count at ART initiation. Follow-up for all clients continued for up to 48 months from the start date through their electronic medical records in the TIER.Net system. We applied administrative censoring for each 12-, 24- and 36-month outcome to ensure clients had sufficient potential follow-up data available to assess outcomes. Clients were censored if the outer window for measuring each outcome went beyond the data extraction date. For assessing the 12-month outcome, for example, clients included in the analysis had to have at least 24 months of potential follow-up data available (= 12-month follow up period + 12-month window for outcomes). Clients starting in 2021 or later had not accrued enough follow-up time for the 24- and 36-month outcomes. These clients were excluded from having their outcomes assessed at 24 and/or 36 months and are denoted as having “insufficient follow-up” for those time points.

### Statistical analysis

We first describe the characteristics of the unique clients in the analytic cohort in the study. Since individual participants could be included in multiple trials, however, our unit of analysis was a trial-client, rather than a unique client. Trial-clients are also described (supplementary files). Once all eight target trials were defined and the cohorts were created, we estimated the risk differences for retention in care and viral suppression comparing those in DSD to those not enrolled in DSD models using a Poisson distribution with an identity link function. We adjusted for age, gender, urban/rural setting, province, WHO stage at ART initiation, and years on ART at trial enrolment. We did not adjust for CD4 count at ART initiation for two reasons: 1) potential collinearity with WHO stage and 2) CD4 count had more missing data compared to WHO stage. We did conduct a sensitivity analysis in which we adjusted for CD4 count at ART initiation in place of WHO stage, and these results are reported in the Supplementary Appendix. We also conducted an age-stratified and sex-stratified analysis to further evaluate the age- and sex-specific impact of DSD on health outcomes. We present risk differences and 95% confidence intervals for each of the eight trials and a pooled estimate that further adjusts for the individual target trial. To account for clients appearing in multiple target trials (resulting in multiple trials for some that would artificially inflate the sample size and bias our standard errors), we also adjusted for within-subject variance using robust variance estimates.

### Ethics approval

The study was approved by the Human Research Ethics Committee (HREC) of the University of the Witwatersrand (M190445) and the Boston University Institutional Review Board (IRB) (H-38115). Both approved use of routine clinic data for the evaluation and provided a waiver of consent.

## Results

### Analytic cohort

A total of 49,595 unique individuals were eligible for DSD model enrolment in our data set. Of these, 52% (25,775) were enrolled in DSD models during the observation period, while 23,820 were eligible but never enrolled. The distribution of gender and WHO stage at ART initiation was similar between those ever enrolled in DSD and those who remained in standard care, with two-thirds (67%) of the clients in WHO stage 1 at baseline and most (69%) female (Table 3). The age and median time on ART at the start of the first trial were higher for those enrolled in DSD (51% aged 35-49 years; median time on ART 4.0 years [interquartile range (IQR) 2.3 to 6.5 years]) compared to those not enrolled in DSD (42% aged 35-49 years; median time on ART 2.5 years [1.5 to 5.3 years]). Clients in the youngest age group (18-24) were less likely to be enrolled in DSD than were older clients. Clients ever enrolled in DSD were slightly more likely to receive care in urban settings than were those never enrolled in DSD (75% vs 69%, respectively) (Table 3). Cohort characteristics based on client trials, rather than unique individuals (available in Supplementary Table S2), are more balanced between DSD and our comparator arms.

**Table 3.**
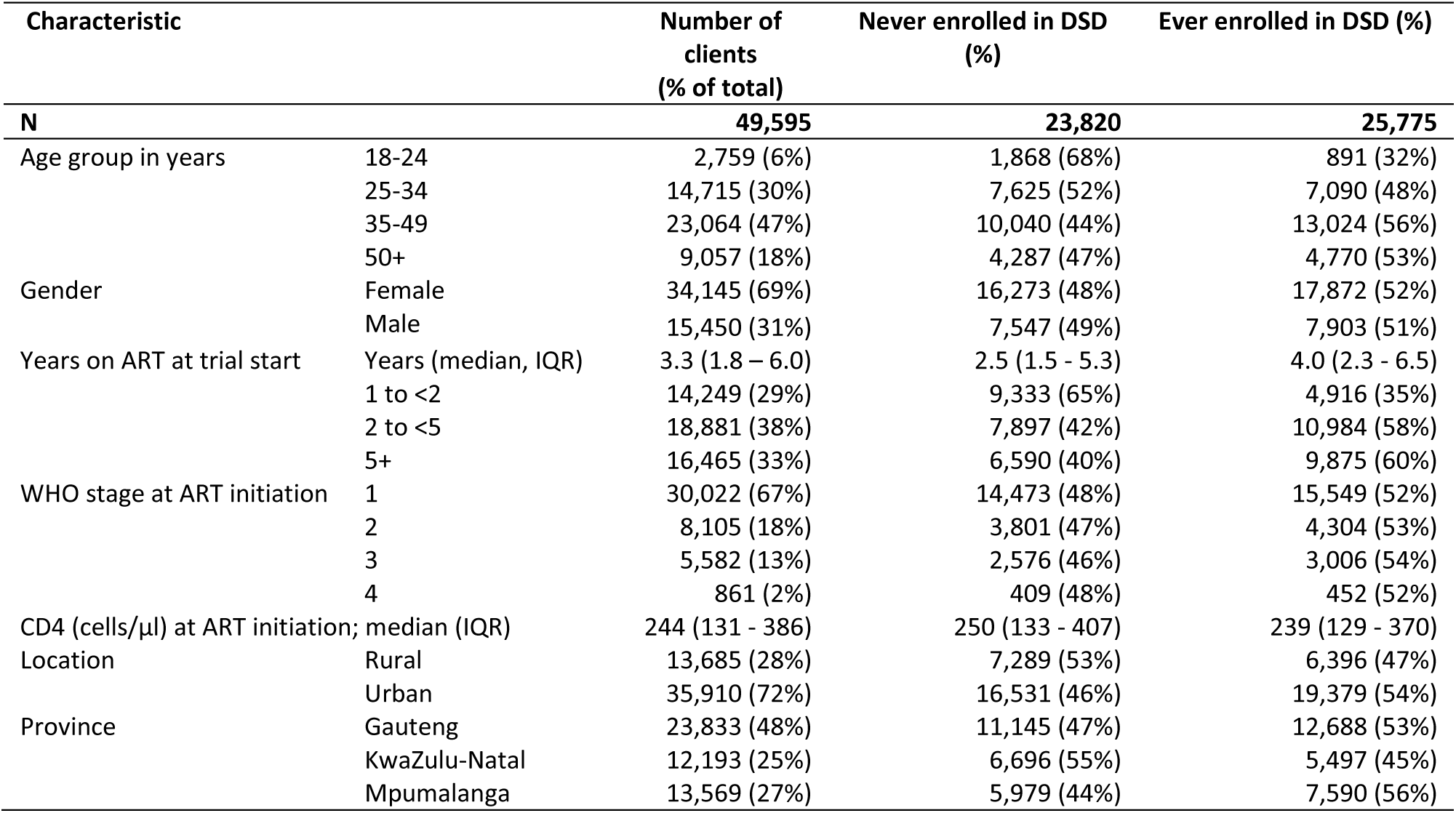
Characteristics of cohort eligible for DSD models in the study.

| Characteristic |  | Number of clients<br>(% of total) | Never enrolled in DSD<br>(%) | Ever enrolled in DSD (%) |
| --- | --- | --- | --- | --- |
| N |  | 49,595 | 23,820 | 25,775 |
| Age group in years | 18-24 | 2,759 (6%) | 1,868 (68%) | 891 (32%) |
|  | 25-34 | 14,715 (30%) | 7,625 (52%) | 7,090 (48%) |
|  | 35-49 | 23,064 (47%) | 10,040 (44%) | 13,024 (56%) |
|  | 50+ | 9,057 (18%) | 4,287 (47%) | 4,770 (53%) |
| Gender | Female | 34,145 (69%) | 16,273 (48%) | 17,872 (52%) |
|  | Male | 15,450 (31%) | 7,547 (49%) | 7,903 (51%) |
| Years on ART at trial start | Years (median, IQR) | 3.3 (1.8 – 6.0) | 2.5 (1.5 - 5.3) | 4.0 (2.3 - 6.5) |
|  | 1 to <2 | 14,249 (29%) | 9,333 (65%) | 4,916 (35%) |
|  | 2 to <5 | 18,881 (38%) | 7,897 (42%) | 10,984 (58%) |
|  | 5+ | 16,465 (33%) | 6,590 (40%) | 9,875 (60%) |
| WHO stage at ART initiation | 1 | 30,022 (67%) | 14,473 (48%) | 15,549 (52%) |
|  | 2 | 8,105 (18%) | 3,801 (47%) | 4,304 (53%) |
|  | 3 | 5,582 (13%) | 2,576 (46%) | 3,006 (54%) |
|  | 4 | 861 (2%) | 409 (48%) | 452 (52%) |
| CD4 (cells/μl) at ART initiation; median (IQR) |  | 244 (131 - 386) | 250 (133 - 407) | 239 (129 - 370) |
| Location | Rural | 13,685 (28%) | 7,289 (53%) | 6,396 (47%) |
|  | Urban | 35,910 (72%) | 16,531 (46%) | 19,379 (54%) |
| Province | Gauteng | 23,833 (48%) | 11,145 (47%) | 12,688 (53%) |
|  | KwaZulu-Natal | 12,193 (25%) | 6,696 (55%) | 5,497 (45%) |
|  | Mpumalanga | 13,569 (27%) | 5,979 (44%) | 7,590 (56%) |

As explained above, because individual participants were included in multiple trials, our unit of analysis was a trial-client, rather than a unique client. 580,276 trial-clients were assessed over the eight target trial enrolment periods between July 2017 and July 2021 (Figure 2). Of all trial-clients, 431,333 were excluded for different reasons: a third (34%, n=200,125) were previously exposed to DSD models, 5% (n=27,791) had not had a recent clinic visit before trial enrolment, 3% (n=18,695) were aged <18 years, 2% (n=9,623) were pregnant during the trial enrolment period, 16% (n=69,136) had <12 months of ART experience, and 25% (n=105,963) were not virally suppressed(Figure 2). A detailed disaggregated flow diagram depicting each of the eight target trials is available in Supplementary Figure S1.

**Figure 2:**
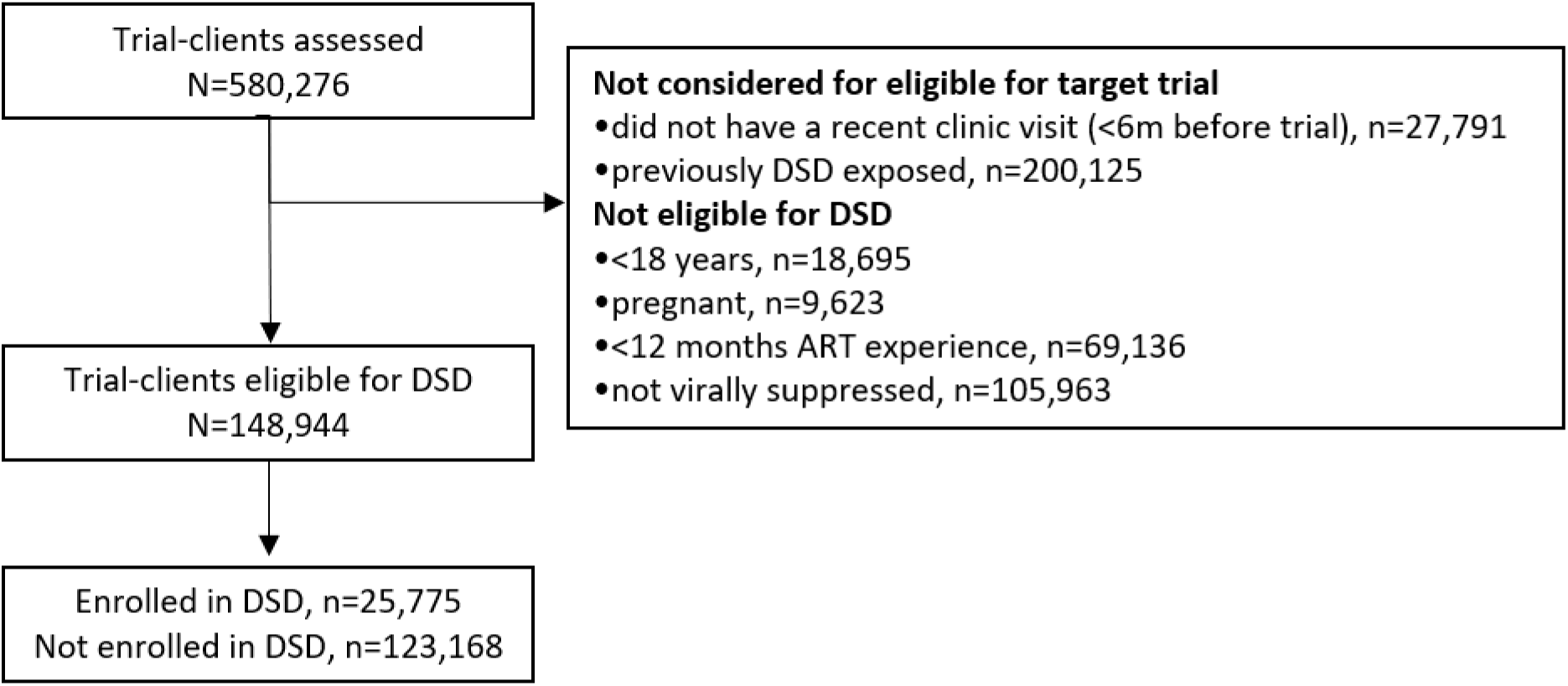
Flow diagram, consolidated across all eight target trials.

### Outcomes

The adjusted mean proportions of participants retained at 12-, 24-, and 36-months post-enrolment period were 92%, 87%, and 82% in the DSD arm, respectively, while proportions retained in the non-DSD comparison arm were 89%, 83%, and 78%, respectively (Figure 3A). The estimated adjusted mean proportions virally suppressed, among those who had viral load results in their medical records, were 95% at all time periods for those enrolled in DSD models and 93%-94% at all time periods for those not enrolled in DSD models (Figure 3B).

**Figure 3:**
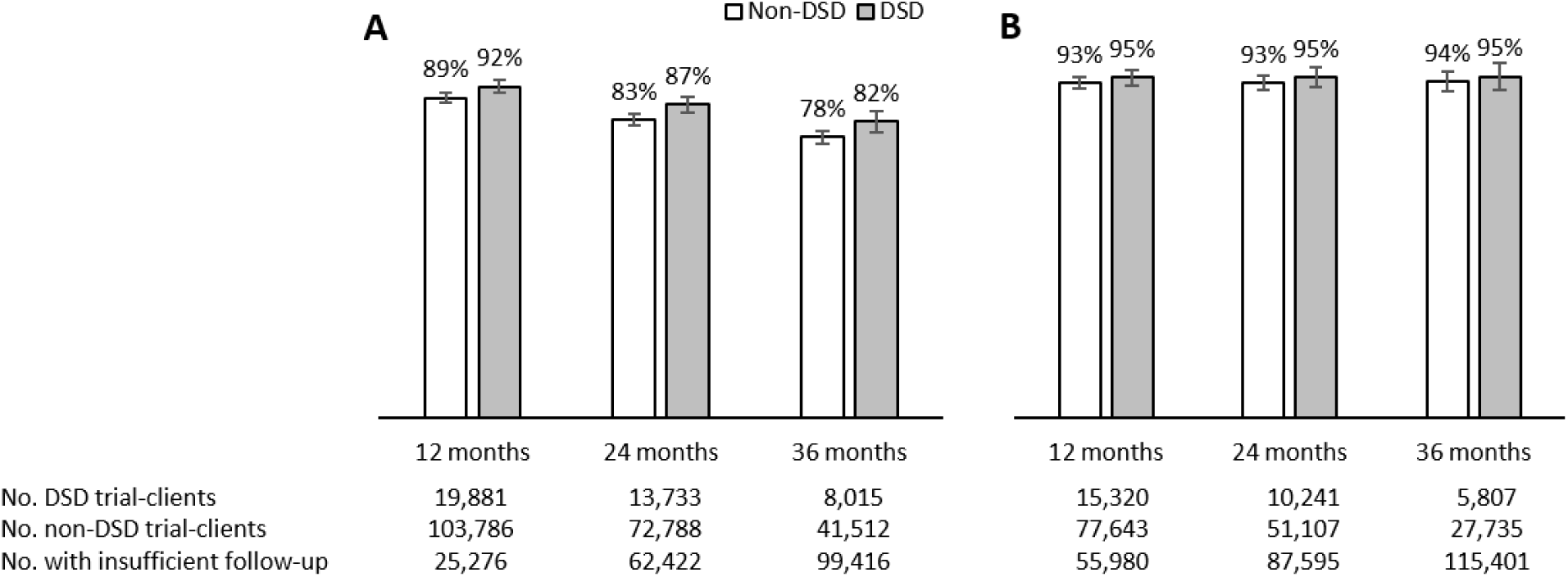
(A) Retention in care and (B) viral load suppression outcomes by DSD enrolment, at 12, 24, and 36 months; adjusted mean estimates with 95% confidence intervals.

The pooled adjusted risk difference for retention in care between clients enrolled in DSD and those eligible but not enrolled was estimated to be 3.2% (95% confidence interval (CI) 1.6%; 4.7%) at 12 months, 4.2% (2.4%; 6.0%) at 24 months, and 4.4% (2.0%; 6.8%) at 36 months (Figure 4). Estimates stratified by each of the target trials were similar, except for the 12-month outcome in the 7^th^ trial which could be explained by timing of the trial enrolment period, which occurred during the first two waves of the COVID-19 pandemic.

**Figure 4:**
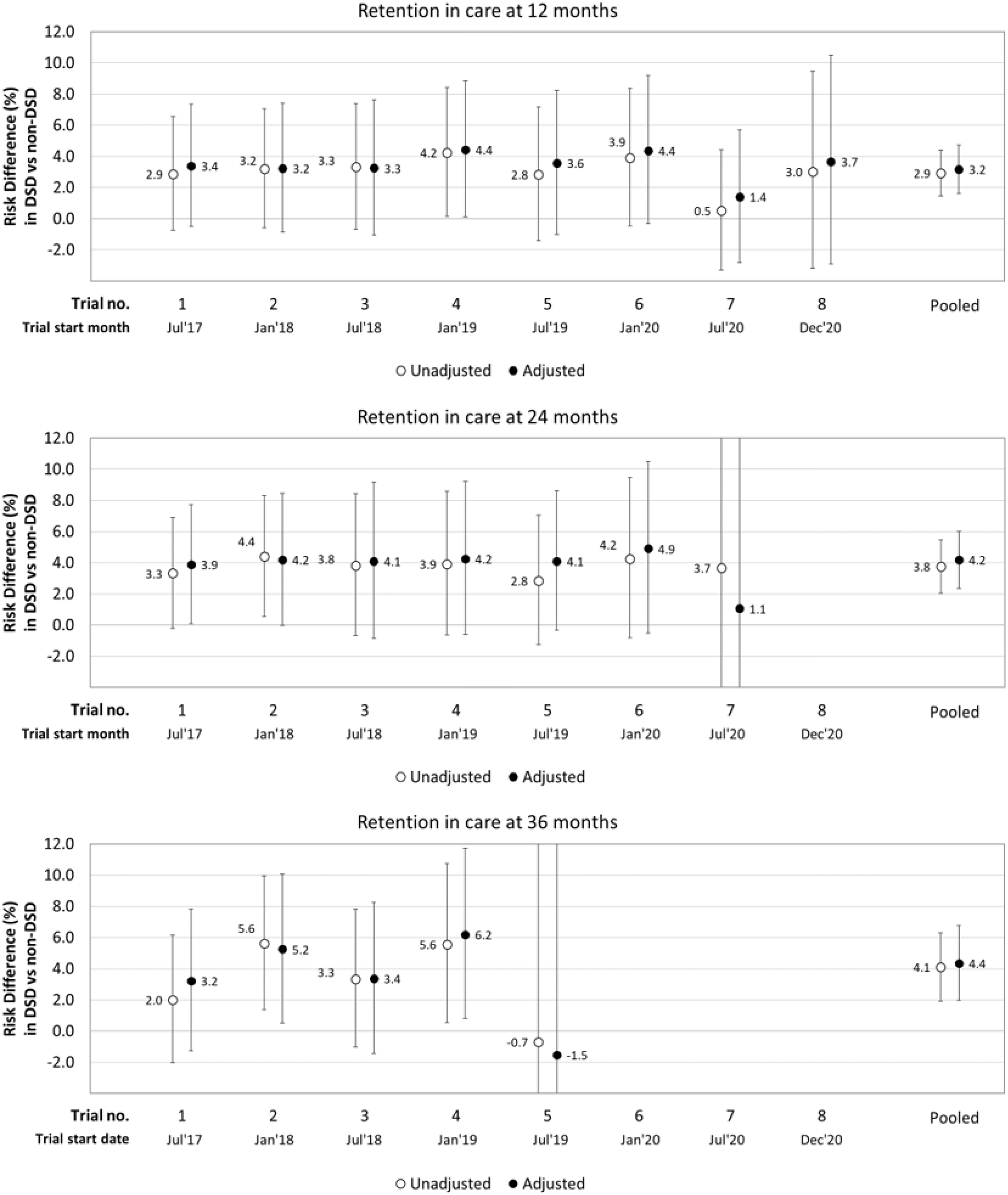
Adjusted risk differences for retention in care (12, 24, and 36 months) comparing DSD vs non-DSD clients.

For viral suppression outcomes, the adjusted risk difference comparing DSD to non-DSD was estimated to be 1.4% (-0.5%; 3.2%) at 12 months, 1.7% (-0.5%; 4.0%) at 24 months, and 1.4% (-0.6%; 4.4%) at 36 months (Figure 5). The pooled adjusted risk differences including the covariate estimates are available in Supplementary Table S3. These show that clients who were younger (aged 18-34 years), received care from an urban clinic, or had fewer years on ART (<2 years) at study enrolment were significantly less likely to be retained in care, while those aged 18-24 years had lower rates of viral suppression at 12 months (Supplementary Table S3). Adjusting for CD4 count at ART initiation, instead of WHO stage, did not fundamentally change the effect estimates for those on DSD models (Supplementary Table S4).

**Figure 5:**
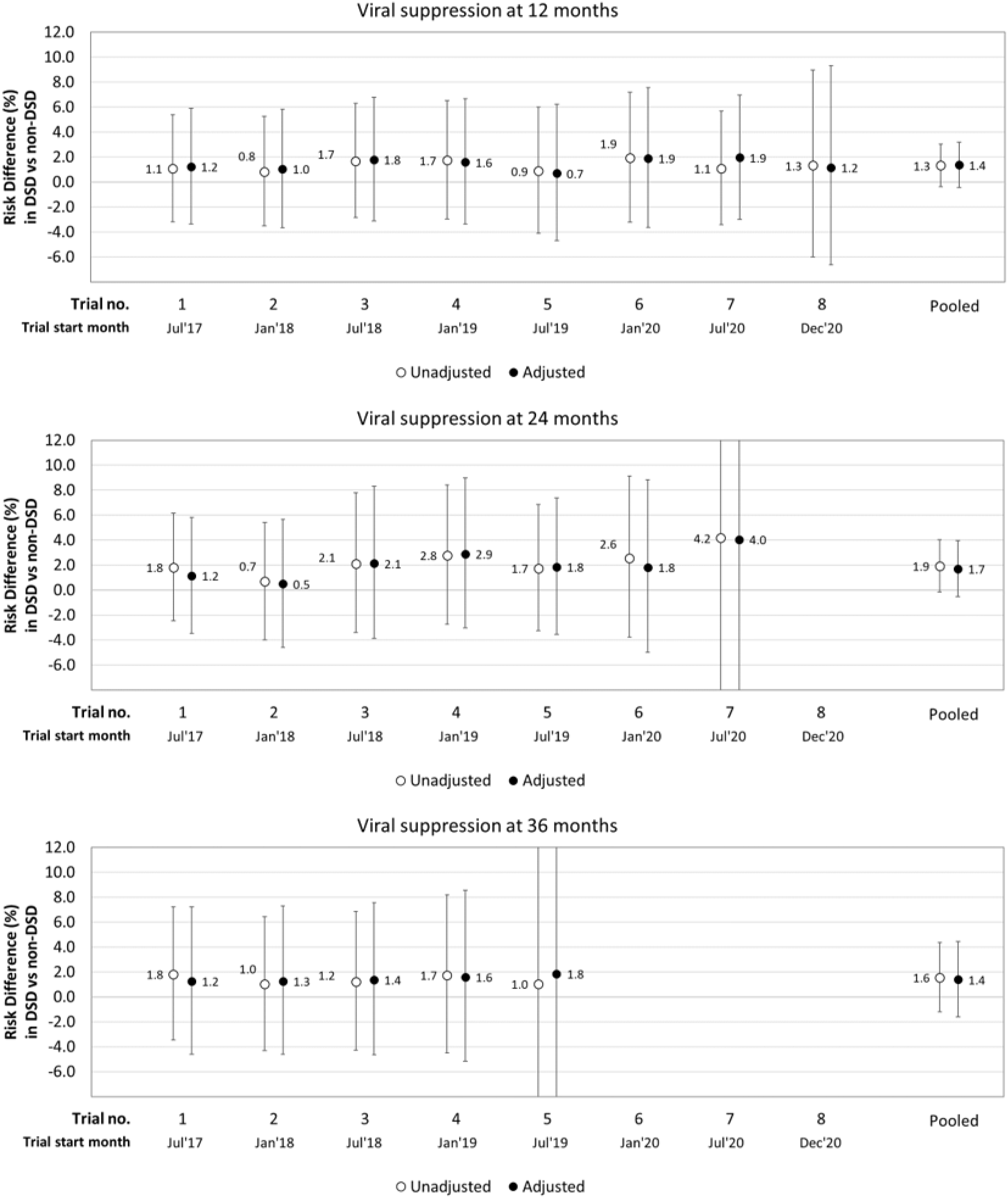
Adjusted risk differences for viral suppression (12, 24, and 36 months) comparing DSD vs non-DSD clients.

Age-stratified analyses showed some variation in risk difference estimates between age groups when comparing DSD vs non-DSD rates of retention and viral suppression. In particular, younger participants (aged 18-24 years) had a higher risk difference for both retention and viral suppression compared to those older (aged 25+ years) (Supplementary Table S5 and S6). Adjusted risk differences for retention ranged from 4.4%-7.8% for 18-24-year-old clients and from 2.5%-5.4% for clients aged 25+ years, while for viral suppression they ranged from 3.3%-6.1% (aged 18-24 years) and from 0.9%-1.7% (aged 25+ years). Confidence intervals were wider for 18-24-year-old clients due to the smaller sample sizes involved, however. Sex-stratified analyses produced similar risk difference estimates between males and females for retention. There were some differences in viral suppression by sex, however, with male clients having a slightly higher adjusted risk difference estimate comparing DSD vs non-DSD (ranging from 1.8%-2.2%) than did female clients (range 1.1%-1.5%) (Supplementary Table S7 and S8).

## Discussion

In this study, we conducted one of the first analyses of a large cohort of clients comparing viral suppression and retention outcomes between those enrolled in differentiated service delivery models and those eligible for DSD enrollment but remaining in conventional care in a routine healthcare setting. We found that clients enrolled in DSD models had similar or slightly better rates of retention in care and viral load suppression as did those who were eligible for DSD but remained in conventional care.

For clients eligible for DSD—i.e. those considered “established” on ART--it is reassuring that retention and suppression outcomes were comparable between conventional care and DSD models. There was little room for these clients’ rate of viral suppression to improve over the follow-up period, as all had documented viral suppression at enrollment. The concern that less intensive models of service delivery, with less direct interaction with clients than under conventional care, might lead to worse retention in care, however, was not borne out in our analysis. The potential benefits of differentiated service delivery, in this setting, can thus be assessed based on non-clinical factors, such as costs to clients, health system resource utilization and efficiency, and client and provider satisfaction.

While countries across sub-Saharan Africa have implemented differentiated models of HIV treatment, few of these programs have undergone rigorous evaluation at scale. Many studies have focused on the impact of individual interventions, often in controlled settings, while less attention has been given to reporting outcomes of these DSD models in routine care settings after national implementation. Despite differences in study designs, our results largely align with the majority of studies on individual models, indicating that less intensive DSD models enrolling clients already established on ART provide equivalent or slightly better retention and viral suppression outcomes [11–17]. Our findings are also consistent with a recent large-scale study assessing the impact of DSD models on 12-month retention in Mozambique[24]. In that study, the research team conducted an interrupted time series analysis comparing clinic-wide retention before and 12 months after the implementation of a package comprising eight models of care. We note that while the question of clinic-wide retention addressed in that analysis differs from the one addressed in our paper, which specifically focuses on the eligible population, both perspectives merit consideration.

Our study had several limitations, most resulting from the observational design and reliance on routinely collected medical records. Although the use of TIER.Net data provided a large sample of ART clients attending multiple healthcare facilities, until June 2020 it did not record the specific DSD model in which a client was enrolled and instead only captured whether the client was in a DSD model or not. As noted above, TIER.Net does not capture off-site medication collection visits for clients enrolled in DSD models, but only the facility-based visits that occur every 6 months, requiring a long follow-up window for retention to be documented. Since the TIER.Net system is not linked across facilities, moreover, “silent” or unrecorded transfers between facilities appear in the system as disengagement from care, likely underestimating overall retention in this study. We cannot know if this effect is differential or non-differential between DSD and comparison clients, however. In addition, in South Africa, the CCMDD program, which provides the majority of medication for DSD models, requires a valid South African identification number, passport number, or asylum seeker number for registration. Anecdotally, a lack of a valid identification number for registration is often offered as a reason for not enrolling clients in the CCMDD program. In clinics with a large number of individuals without these numbers, such as those that serve immigrant communities, facilities may decide to pack medications themselves, at the facility, rather than relying on the CCMDD program. This could explain why some clients who appear in their medical records to be eligible for DSD are not enrolled.

The target trial emulation methodology used in this analysis is a robust method for defining a comparator arm and determining outcomes in routine care data but it cannot correct all issues that affect observational studies. It can help prevent issues like misaligned person time, poorly defined eligibility criteria, and unclear causal questions. It does not fully address the limitations of observational studies, however, as residual confounding can still be present. In our case, it is likely that many of those who were offered and accepted enrollment in DSD models were, on average, in better health, more motivated, or more empowered to remain in care than those who were not, suggesting that DSD participants may have had better outcomes than comparison participants even in the absence of DSD. This bias would overestimate the retention benefit of DSD over conventional care. The exclusion from DSD of otherwise-eligible clients without national identification numbers could also skew our results, either in favor of no difference (if clients without ID numbers also have good outcomes in conventional care, offsetting the bias mentioned above) or by overestimating the effect of DSD, if clients without ID numbers face greater barriers to remaining in care.

Despite these study limitations, our results using a robust observational methodology and large sample size suggest that retention and viral suppression for those in DSD models in South Africa are similar to, or better than, outcomes in conventional care for ART clients who meet typical DSD eligibility criteria. These findings should assure policy makers and program managers that the less intensive models of care that South Africa has introduced do not threaten the achievements of national HIV programs and that the non-clinical benefits they generate for the healthcare system, such as savings in provider time use, and for clients, such as less time and lower costs, will not be offset by poorer clinical outcomes.

## Data Availability

Data used in this study are owned by the South Africa National Department of Health and cannot be shared by the authors.

## Authors’ contributions

Conceptualization: Amy Huber, Lise Jamieson, Matthew Fox, Brooke Nichols, Sydney Rosen

Data curation: Amy Huber, Lise Jamieson

Formal analysis: Amy Huber, Lise Jamieson, Matthew Fox

Funding acquisition: Sophie Pascoe, Sydney Rosen

Methodology: Matthew Fox, Lise Jamieson

Project administration: Amy Huber, Sophie Pascoe

Validation: Lufuno Malala, Musa Manganye, Thato Chidarikire, Nthabiseng Khoza

Visualization: n/a

Writing – original draft: Amy Huber, Lise Jamieson, Sydney Rosen

Writing – review & editing: Amy Huber, Lise Jamieson, Matthew P. Fox, Musa Manganye, Lufuno Malala, Thato Chidarikire, Nthabiseng Khoza, Brooke Nichols, Sydney Rosen, Sophie Pascoe

## Supplementary Appendix

**Table S1.**
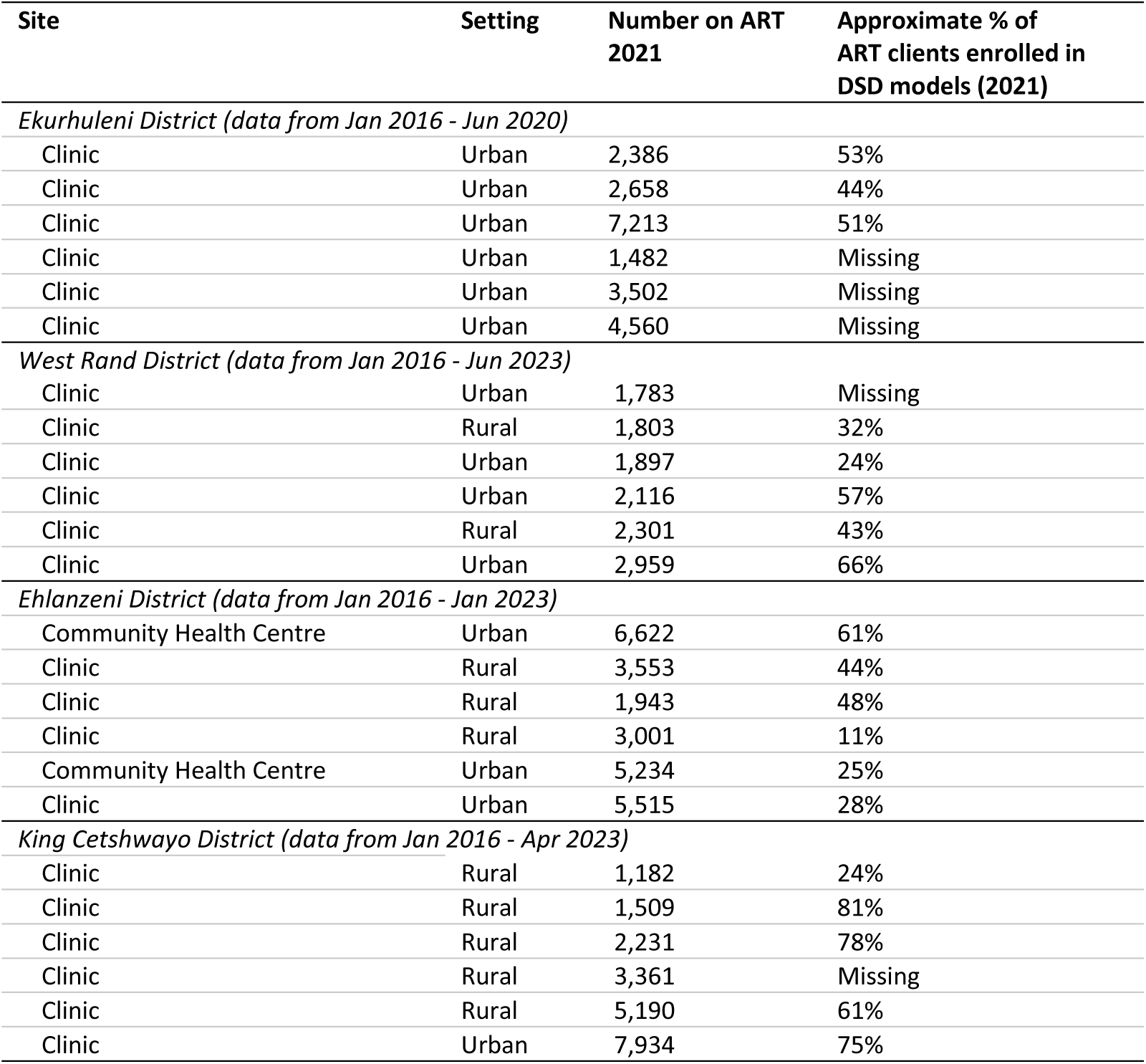
Study site description.

| Site | Setting | Number on ART 2021 | Approximate % of ART clients enrolled in DSD models (2021) |
| --- | --- | --- | --- |
| *Ekurhuleni District (data from Jan 2016 - Jun 2020)* |  |  |  |
| Clinic | Urban | 2,386 | 53% |
| Clinic | Urban | 2,658 | 44% |
| Clinic | Urban | 7,213 | 51% |
| Clinic | Urban | 1,482 | Missing |
| Clinic | Urban | 3,502 | Missing |
| Clinic | Urban | 4,560 | Missing |
| *West Rand District (data from Jan 2016 - Jun 2023)* |  |  |  |
| Clinic | Urban | 1,783 | Missing |
| Clinic | Rural | 1,803 | 32% |
| Clinic | Urban | 1,897 | 24% |
| Clinic | Urban | 2,116 | 57% |
| Clinic | Rural | 2,301 | 43% |
| Clinic | Urban | 2,959 | 66% |
| *Ehlanzeni District (data from Jan 2016 - Jan 2023)* |  |  |  |
| Community Health Centre | Urban | 6,622 | 61% |
| Clinic | Rural | 3,553 | 44% |
| Clinic | Rural | 1,943 | 48% |
| Clinic | Rural | 3,001 | 11% |
| Community Health Centre | Urban | 5,234 | 25% |
| Clinic | Urban | 5,515 | 28% |
| *King Cetshwayo District (data from Jan 2016 - Apr 2023)* |  |  |  |
| Clinic | Rural | 1,182 | 24% |
| Clinic | Rural | 1,509 | 81% |
| Clinic | Rural | 2,231 | 78% |
| Clinic | Rural | 3,361 | Missing |
| Clinic | Rural | 5,190 | 61% |
| Clinic | Urban | 7,934 | 75% |

**Figure S1.**
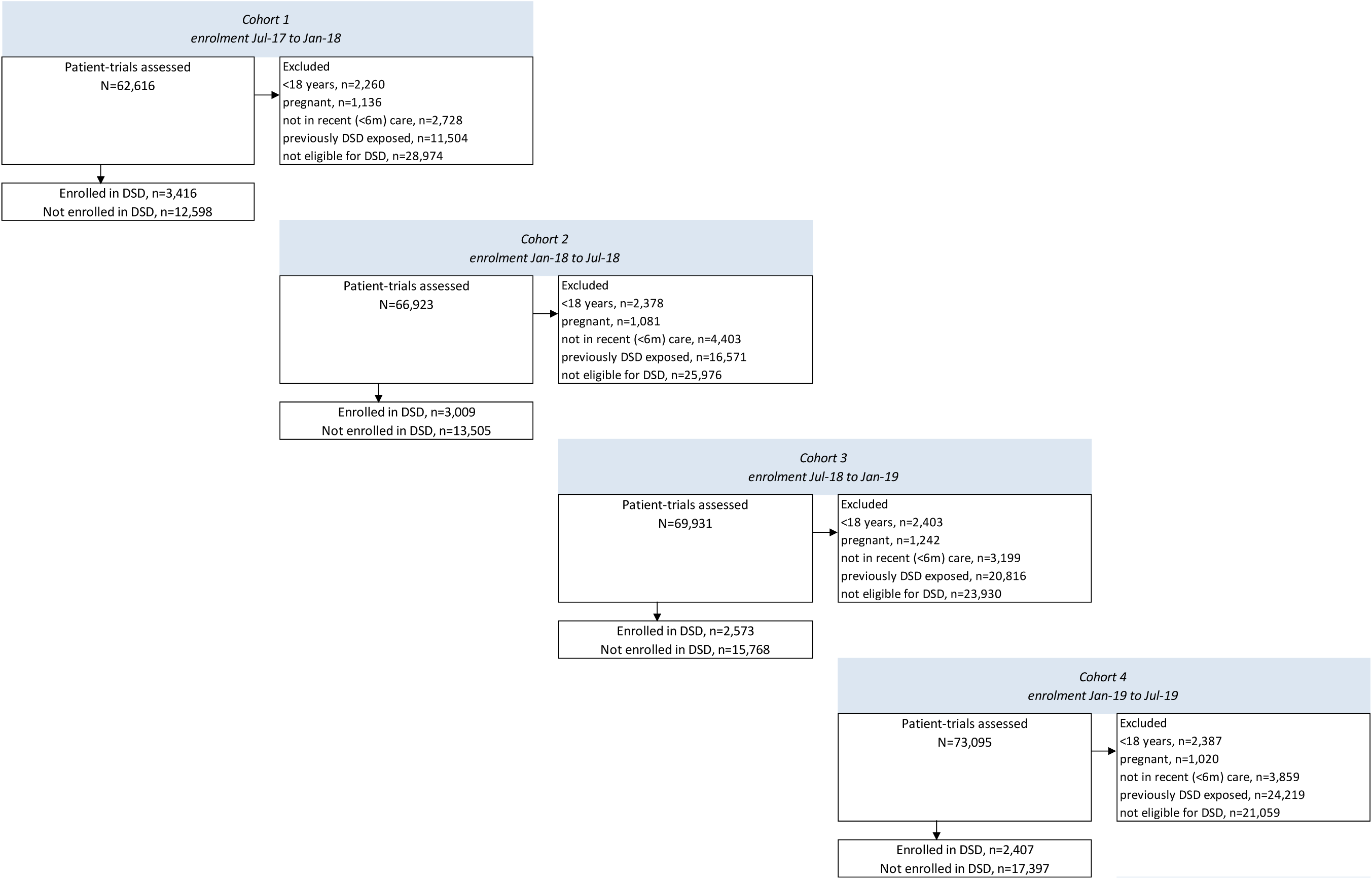
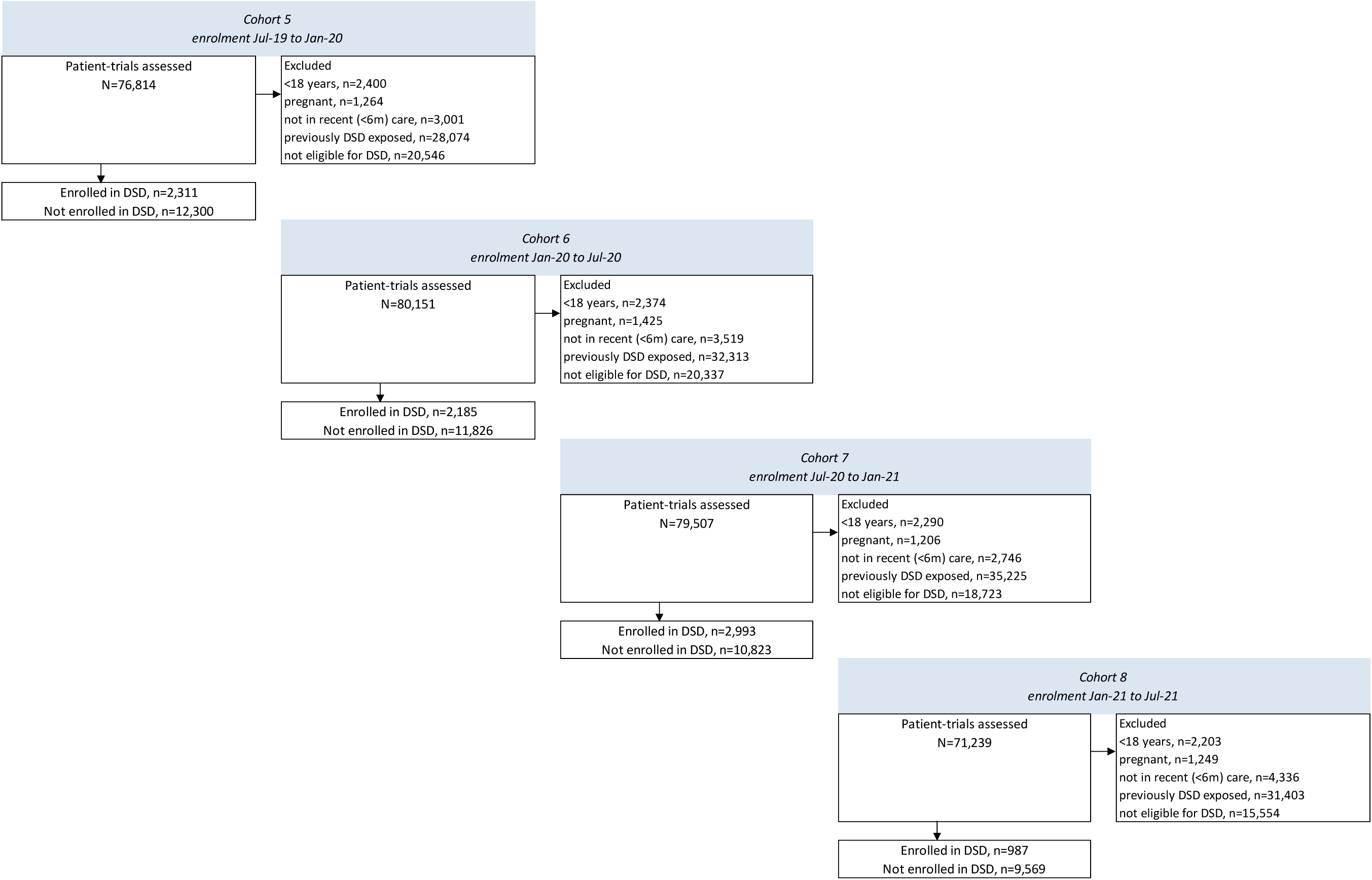
Cohort flow diagram.

**Table S2.** Baseline demographics of trial-clients eligible for DSD models across all eight emulated target trials.

|  |  | Total (%) | Not enrolled in DSD (%) | Enrolled in DSD (%) |
| --- | --- | --- | --- | --- |
| **Number of trial-clients \*** |  | **148,943** | **123,168** | **25,775** |
| Age group in years | 18-24 | 6,664 (4%) | 5,773 (87%) | 891 (13%) |
|  | 25-34 | 40,063 (27%) | 32,973 (82%) | 7,090 (18%) |
|  | 35-49 | 70,700 (47%) | 57,676 (82%) | 13,024 (18%) |
|  | 50+ | 31,516 (21%) | 26,746 (85%) | 4,770 (15%) |
| Gender | Female | 102,680 (69%) | 84,808 (83%) | 17,872 (17%) |
|  | Male | 46,263 (31%) | 38,360 (83%) | 7,903 (17%) |
| Years on ART at trial start, median (IQR) |  | 4.1 (2.4 - 6.7) | 4.1 (2.4 - 6.7) | 4.0 (2.3 - 6.5) |
| Years on ART at trial start | 1-<2 | 26,590 (18%) | 21,674 (82%) | 4,916 (18%) |
|  | 2-<5 | 62,539 (42%) | 51,555 (82%) | 10,984 (18%) |
|  | 5+ | 59,814 (40%) | 49,939 (83%) | 9,875 (17%) |
| WHO stage at ART initiation | 1 | 86,361 (65%) | 70,812 (82%) | 15,549 (18%) |
|  | 2 | 25,732 (19%) | 21,428 (83%) | 4,304 (17%) |
|  | 3 | 17,901 (13%) | 14,895 (83%) | 3,006 (17%) |
|  | 4 | 2,842 (2%) | 2,390 (84%) | 452 (16%) |
| CD4 (cells/μl) at ART initiation; median (IQR) | | 238 (128 - 371) | 237 (128 - 372) | 239 (129 - 370) |
| Location | Rural | 42,292 (28%) | 35,896 (85%) | 6,396 (15%) |
|  | Urban | 106,651 (72%) | 87,272 (82%) | 19,379 (18%) |
| Province | Gauteng | 71,265 (48%) | 58,577 (82%) | 12,688 (18%) |
|  | KwaZulu-Natal | 37,327 (25%) | 31,830 (85%) | 5,497 (15%) |
|  | Mpumalanga | 40,351 (27%) | 32,761 (81%) | 7,590 (19%) |

\* trial-clients refers to the number of clients followed across all 8 target trials, with some clients appearing in multiple target trials

**Table S3.**
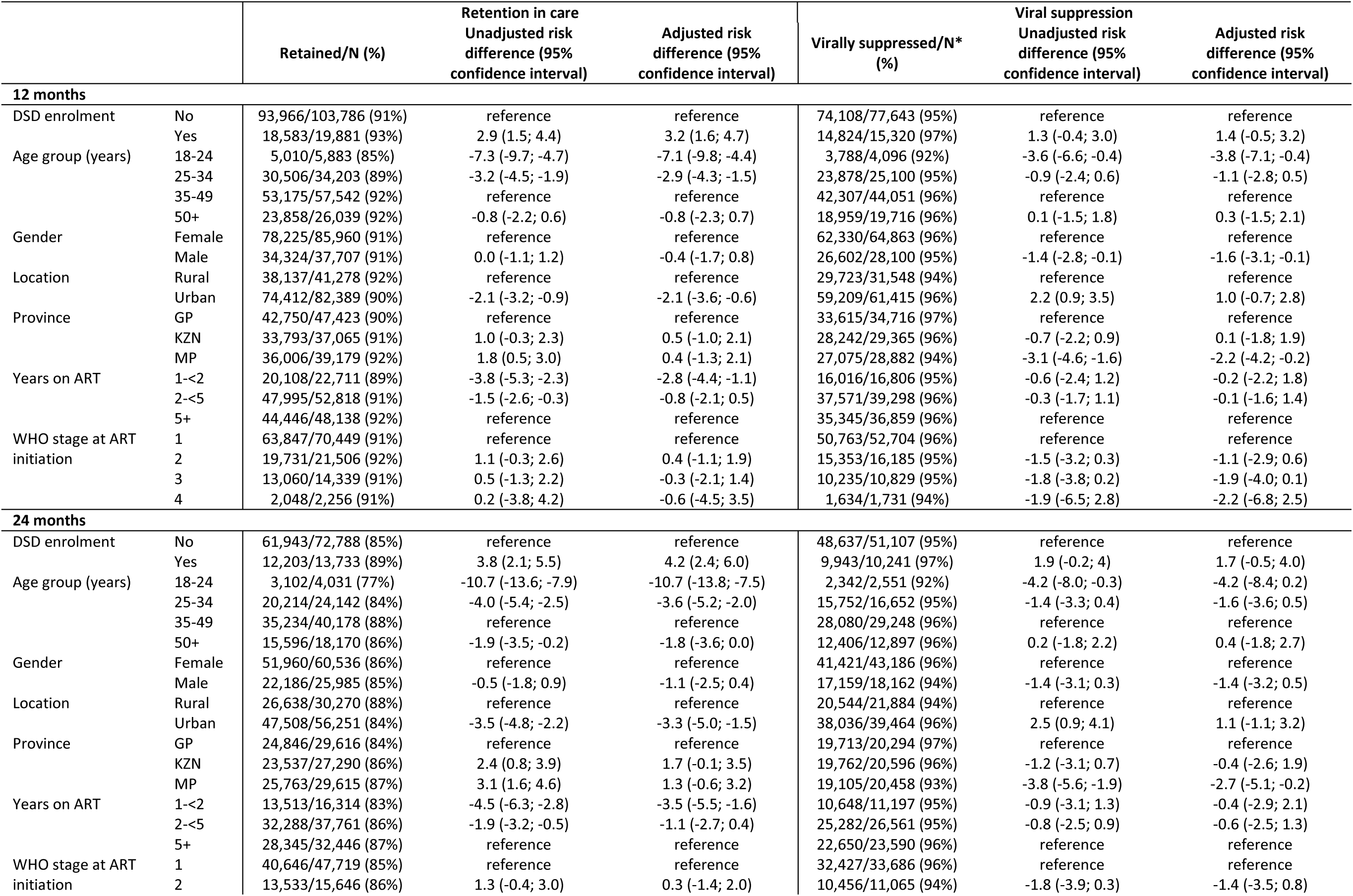
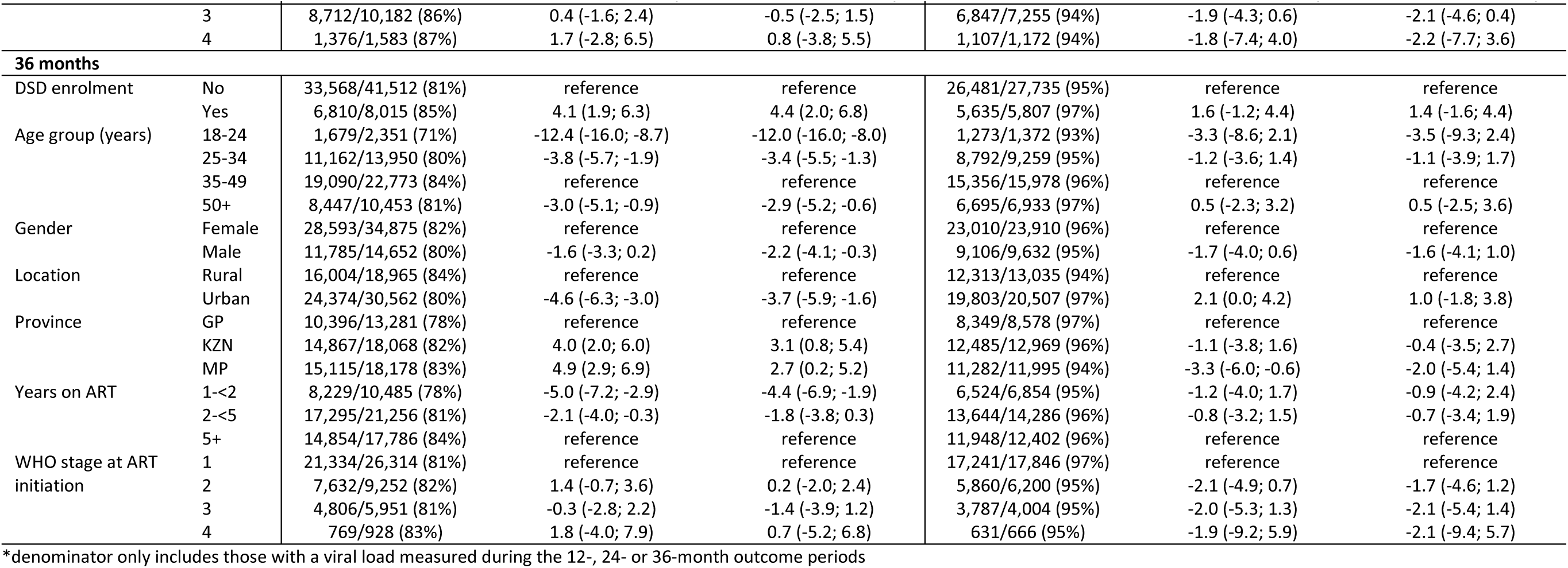
Pooled adjusted risk difference (%) of retention in care and viral suppression by DSD enrolment at 12, 24 and 36 months.

**Table S4.**
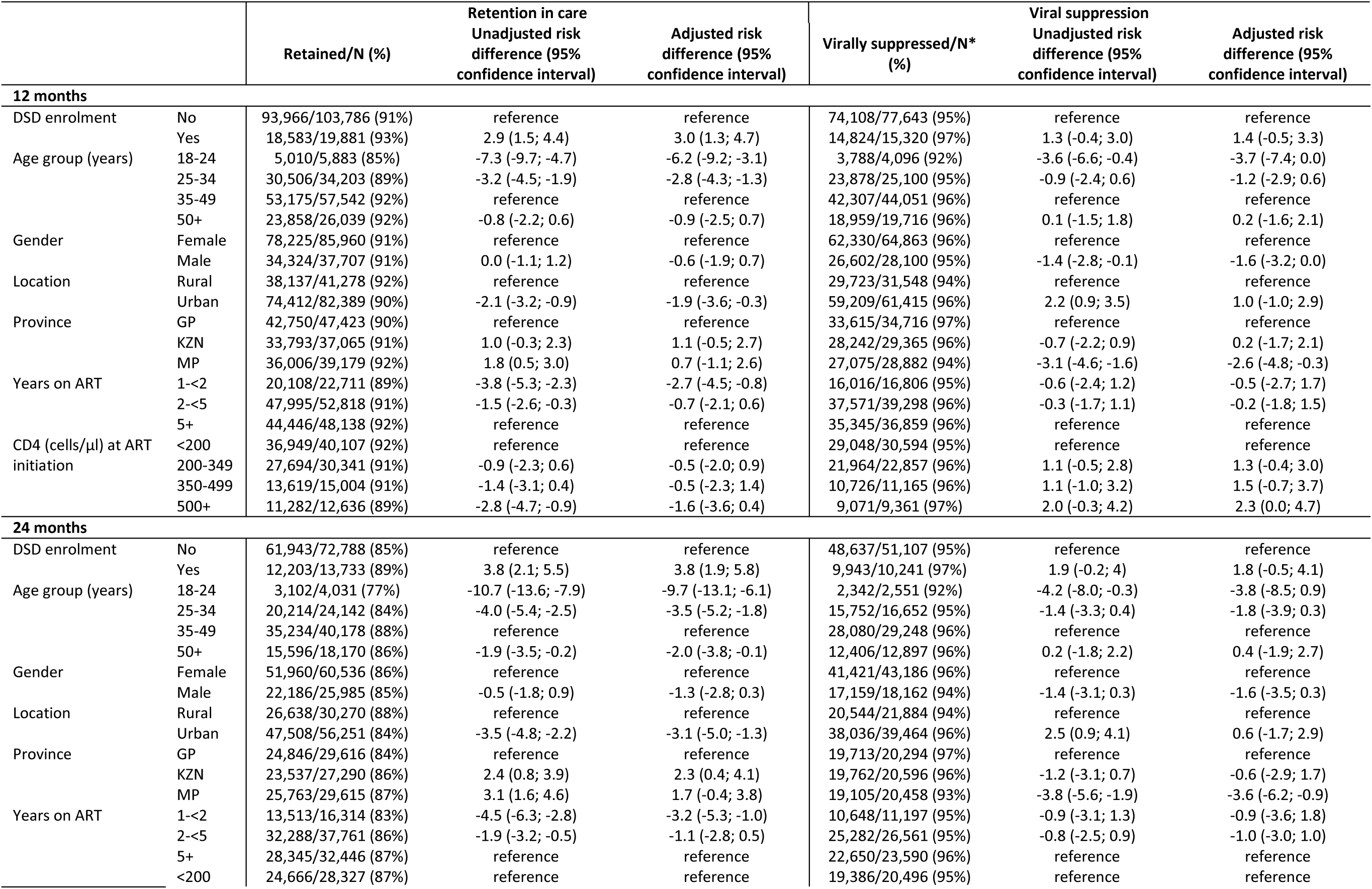
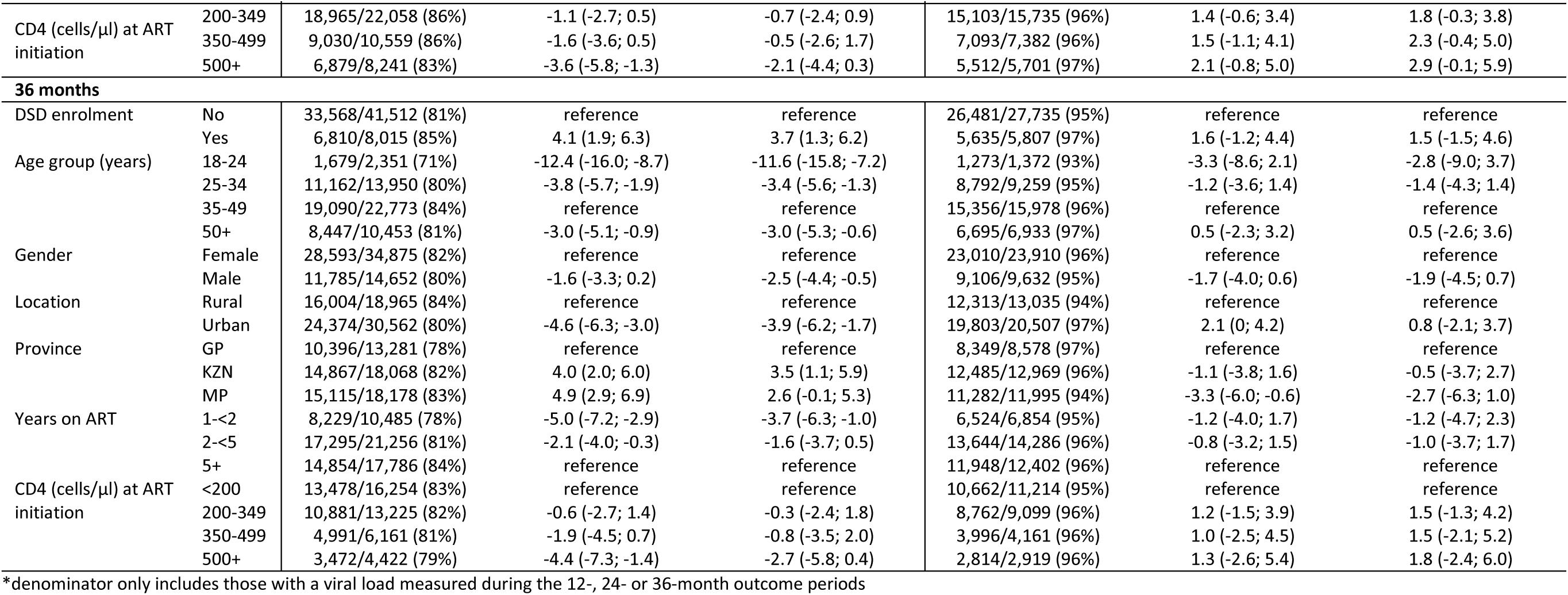
Pooled adjusted risk difference (%) of retention in care and viral suppression by DSD enrolment at 12, 24 and 36 months, adjusting for CD4 count at ART initiation.

**Table S5.** Age-stratified pooled risk differences for retention.

| Age group | n/N (%) retained<br>in DSD | n/N (%) retained<br>in non-DSD | Unadjusted Risk Difference<br>comparing DSD vs non-DSD<br>(95% CI) | Adjusted Risk Difference<br>comparing DSD vs non-DSD<br>(95% CI) |
| --- | --- | --- | --- | --- |
| **12 months** |  |  |  |  |
| 18-24 | 660/745 (89%) | 4,350/5,138 (85%) | 3.9 (-3.1; 11.3) | 4.4 (-3.2; 12.3) |
| 25-34 | 5,268/5,713 (92%) | 25,238/28,491 (89%) | 3.6 (0.9; 6.4) | 4.1 (1.3; 7.0) |
| 35-49 | 9,241/9,797 (94%) | 43,934/47,745 (92%) | 2.3 (0.2; 4.4) | 2.5 (0.3; 4.8) |
| 50+ | 3,414/3,627 (94%) | 20,444/22,412 (91%) | 2.9 (-0.5; 6.3) | 3.0 (-0.6; 6.7) |
| **24 months** |  |  |  |  |
| 18-24 | 422/505 (84%) | 2,680/3,526 (76%) | 7.6 (-0.7; 16.3) | 7.8 (-1.2; 17.2) |
| 25-34 | 3,469/3,990 (87%) | 16,745/20,153 (83%) | 3.9 (0.7; 7.0) | 4.7 (1.4; 8.1) |
| 35-49 | 6,087/6,743 (90%) | 29,147/33,435 (87%) | 3.1 (0.6; 5.6) | 3.5 (0.9; 6.2) |
| 50+ | 2,225/2,496 (89%) | 13,371/15,674 (85%) | 3.8 (-0.1; 7.9) | 4.1 (-0.1; 8.5) |
| **36 months** |  |  |  |  |
| 18-24 | 234/301 (78%) | 1,445/2,050 (70%) | 7.3 (-3.0; 18.2) | 7.0 (-4.2; 19.0) |
| 25-34 | 1,982/2,400 (83%) | 9,180/11,550 (79%) | 3.1 (-0.8; 7.1) | 3.6 (-0.6; 8.0) |
| 35-49 | 3,381/3,892 (87%) | 15,709/18,881 (83%) | 3.7 (0.5; 6.9) | 4.2 (0.7; 7.8) |
| 50+ | 1,213/1,422 (85%) | 7,234/9,031 (80%) | 5.2 (0.1; 10.4) | 5.4 (-0.1; 11.1) |

**Table S6.** Age-stratified pooled risk differences for viral suppression.

| Age group | n/N (%) virally suppressed* in DSD | n/N (%) virally suppressed in non-DSD | Unadjusted Risk Difference comparing DSD vs non-DSD (95% CI) | Adjusted Risk Difference comparing DSD vs non-DSD (95% CI) |
| --- | --- | --- | --- | --- |
| **12 months** |  |  |  |  |
| 18-24 | 531/555 (96%) | 3,257/3,541 (92%) | 3.7 (-4.8; 12.6) | 3.3 (-5.9; 12.9) |
| 25-34 | 4,171/4,335 (96%) | 19,706/20,765 (95%) | 1.3 (-1.9; 4.5) | 1.4 (-2.0; 4.8) |
| 35-49 | 7,404/7,627 (97%) | 34,901/36,424 (96%) | 1.3 (-1.2; 3.7) | 1.2 (-1.3; 3.8) |
| 50+ | 2,717/2,803 (97%) | 16,236/16,913 (96%) | 0.9 (-3.0; 4.9) | 1.3 (-2.9; 5.6) |
| **24 months** |  |  |  |  |
| 18-24 | 329/347 (95%) | 2,013/2,204 (91%) | 3.5 (-7.2; 14.8) | 4.1 (-7.6; 16.6) |
| 25-34 | 2,780/2,891 (96%) | 12,972/13,762 (94%) | 1.9 (-2.0; 5.9) | 1.3 (-2.8; 5.6) |
| 35-49 | 5,002/5,129 (98%) | 23,075/24,119 (96%) | 1.9 (-1.1; 4.8) | 1.7 (-1.5; 4.9) |
| 50+ | 1,829/1,874 (98%) | 10,575/11,023 (96%) | 1.7 (-3.1; 6.6) | 1.6 (-3.5; 6.9) |
| **36 months** |  |  |  |  |
| 18-24 | 196/200 (98%) | 1,077/1,172 (92%) | 6.1 (-8.1; 21.4) | 6.1 (-9.7; 23.3) |
| 25-34 | 1,629/1,688 (97%) | 7,161/7,571 (95%) | 1.9 (-3.2; 7.2) | 1.6 (-3.8; 7.3) |
| 35-49 | 2,811/2,900 (97%) | 12,543/13,078 (96%) | 1.0 (-2.9; 5.0) | 0.9 (-3.4; 5.2) |
| 50+ | 999/1,019 (98%) | 5,695/5,914 (96%) | 1.7 (-4.7; 8.4) | 1.6 (-5.4; 8.9) |

\*Viral suppression defined as having a viral load of <400 copies/ml

**Table S7.** Sex-stratified pooled risk differences for viral suppression.

| Age group | n/N (%) retained in DSD | n/N (%) retained in non-DSD | Unadjusted Risk Difference comparing DSD vs non-DSD (95% CI) | Adjusted Risk Difference comparing DSD vs non-DSD (95% CI) |
| --- | --- | --- | --- | --- |
| **12 months** |  |  |  |  |
| Female | 13,090/13,991 (94%) | 65,135/71,970 (91%) | 3.1 (1.3; 4.8) | 3.2 (1.3; 5.0) |
| Male | 5,493/5,891 (93%) | 28,831/31,816 (91%) | 2.6 (0.0; 5.3) | 3.1 (0.3; 6.0) |
| **24 months** |  |  |  |  |
| Female | 8,678/9,755 (89%) | 43,282/50,782 (85%) | 3.7 (1.7; 5.8) | 4.1 (1.9; 6.3) |
| Male | 3,525/3,979 (89%) | 18,661/22,006 (85%) | 3.8 (0.6; 7.0) | 4.3 (1.0; 7.8) |
| **36 months** |  |  |  |  |
| Female | 4,924/5,756 (86%) | 23,669/29,119 (81%) | 4.3 (1.7; 6.9) | 4.4 (1.5; 7.2) |
| Male | 1,886/2,259 (83%) | 9,899/12,393 (80%) | 3.6 (-0.4; 7.7) | 4.4 (0.0; 8.9) |

**Table S8.** Sex-stratified pooled risk differences for viral suppression.

| Age group | n/N (%) virally suppressed in DSD | n/N (%) virally suppressed in non-DSD | Unadjusted Risk Difference comparing DSD vs non-DSD (95% CI) | Adjusted Risk Difference comparing DSD vs non-DSD (95% CI) |
| --- | --- | --- | --- | --- |
| **12 months** |  |  |  |  |
| Female | 10,532/10,861 (97%) | 51,790/54,002 (96%) | 1.1 (-1.0; 3.1) | 1.1 (-1.0; 3.3) |
| Male | 4,291/4,459 (96%) | 22,310/23,641 (94%) | 1.9 (-1.2; 5.0) | 1.8 (-1.5; 5.2) |
| **24 months** |  |  |  |  |
| Female | 7,106/7,307 (97%) | 34,313/35,880 (96%) | 1.6 (-0.8; 4.1) | 1.5 (-1.2; 4.1) |
| Male | 2,834/2,934 (97%) | 14,322/15,228 (94%) | 2.5 (-1.3; 6.5) | 2.2 (-1.9; 6.3) |
| **36 months** |  |  |  |  |
| Female | 4,119/4,237 (97%) | 18,889/19,673 (96%) | 1.2 (-2.0; 4.5) | 1.2 (-2.3; 4.8) |
| Male | 1,516/1,570 (97%) | 7,587/8,062 (94%) | 2.5 (-2.8; 7.8) | 1.9 (-3.8; 7.7) |

